# Neurotransmitter-specific polygenic scores dissect clinical heterogeneity in schizophrenia

**DOI:** 10.64898/2026.11.05.26367352

**Authors:** Pol Ramon-Cañellas, Javier Vázquez-Bourgon, Benedicto Crespo-Facorro, María Dolores Moltó, Maria José Escartí, Javier González-Peñas, Celso Arango, Ana González-Pinto, Lourdes Fañanás, Barbara Arias, Julio Bobes, Javier Costas, Lourdes Martorell, Elisabet Vilella, Gabriel Santpere, Gerard Muntané

## Abstract

Schizophrenia (SCZ) is a highly heritable psychiatric disorder characterised by substantial clinical heterogeneity. While global polygenic scores (gPGS) capture genetic liability to SCZ, their ability to explain clinical variability remains limited. Pathway-stratified polygenic scores (sPGS) offer a biologically informed approach to patient stratification.

In this study, common genetic variants were annotated across five neurotransmitter (NT) signalling pathways — acetylcholine (ACh), dopamine, γ-aminobutyric acid (GABA), glutamate (Glu), and serotonin. sPGS adjusted for sex and ancestry principal components, were calculated in two independent cohorts with different available clinical data (N=3,044 and N=645). By clustering top-decile individuals across the five sPGS dimensions, we classified patients into five mutually exclusive patient profiles, which were then evaluated for their associations with Positive and Negative Syndrome Scale (PANSS) symptom dimensions, age at disease onset (AAO), and three-year clinical remission and treatment response.

Although gPGS showed no significant association with clinical outcomes, some sPGS-derived profiles captured clinically relevant heterogeneity across cohorts. Patients assigned to the Ach profile exhibited higher overall symptom severity, particularly negative symptoms and general psychopathology. Patients within the Glu profile were characterised by more severe negative symptoms, earlier AAO, reduced three-year remission, and poorer treatment response. Similarly, the GABA risk profile was also associated with three-year remission rates.

These findings suggest that NT pathway-specific polygenic profiling may capture clinically relevant variation overlooked by genome-wide approaches. By identifying genetic profiles associated with symptom severity, disease onset, and longitudinal outcomes, this framework could contribute to biologically informed patient stratification and inform future precision psychiatry approaches in SCZ.

## Introduction

Schizophrenia (SCZ) is a complex neurodevelopmental disorder affecting 1% of worldwide population, that presents a public health challenge, in part, due to its variable clinical trajectory and highly heterogeneous treatment outcomes. SCZ has a substantial genetic component with twin-based heritability between 60% to 80%, while landmark genome-wide association studies (GWAS) have mapped hundreds of common-variant loci, establishing an overall SNP-based heritability of 20% [1]. However, a major hurdle for clinical translation remains: while global polygenic scores (gPGS) capture part of the genetic liability to distinguish cases from healthy controls (HC), their ability to explain clinical variability among affected individuals remains limited [2–5]. This predictive disconnection presents a challenge to clinical psychiatry, as broad diagnostic categories group divergent biological trajectories under a single clinical label [6].

Recent research has increasingly focused on dimensional phenotypes and longitudinal clinical outcomes rather than binary diagnostic classifications [7, 8]. Genetic liability to SCZ has been associated with cognitive traits, including general cognitive performance, working memory, and processing speed, in patients with SCZ, clinically at-risk individuals, and even in individuals without a diagnosis [9, 10]. However, gPGS generally explain only a small proportion of the variance in these quantitative phenotypes, providing limited information on the clinical heterogeneity observed. Among patients with SCZ, clinical characteristics such as age at onset (AAO), symptom severity, treatment response, and long-term remission vary considerably between individuals, potentially reflecting differences in the biological processes contributing to disease expression and progression [2, 6].

Genetic evidence consistently implicates neuronal and synaptic biology in SCZ. Common risk variants are predominantly enriched in genes expressed in mature cortical and subcortical neurons [11–13], converging on biological processes such as neuronal development, ion channel activity, and synaptic function [1, 14]. This synaptic convergence is further reinforced by rare copy-number variants and rare damaging coding mutations, which disproportionately disrupt synaptic genes, including neurotransmitter (NT) receptors and signalling molecules [15, 16]. Together, these findings place synaptic and NT signalling among the major biological substrates of SCZ genetic liability.

Accordingly, altered neurotransmission has long been considered a central mechanism underlying SCZ pathophysiology and remains the primary target of current psychopharmacological interventions [17–19]. Although hyperdopaminergic signalling has historically dominated neurobiological models and first-generation drug development, converging evidence from genetic, neuroimaging, and pharmacological studies supports the involvement of multiple NT systems, including acetylcholine (ACh), γ-aminobutyric acid (GABA), glutamate (Glu), serotonin (5-HT), in addition to dopamine (DA) [20–24]. Localised dysfunction within these signalling cascades maps to specific clinical manifestations of the disorder, including symptom severity, cognitive impairment, AAO, and treatment response [22, 25–28]. This multi-system involvement is reflected pharmacologically, as second-, and third-generation antipsychotics exert therapeutic effects through complex multi-receptor binding profiles rather than a single signalling pathway [29, 30]. These observations raise the possibility that aggregating genetic liability into a single genome-wide PGS may obscure biologically distinct components of SCZ risk. Decomposing polygenic liability according to specific NT systems could therefore provide a means to identify genetic profiles associated with distinct clinical trajectories [2, 31].

Our study systematically deconstructed overall common-variant liability into five independent, pathway-specific PGS targeting the ACh, DA, GABA, Glu, and 5-HT signalling cascades. We mapped these neurochemical risk profiles against granular clinical outcomes, evaluating Positive and Negative Syndrome Scale (PANSS) subscales, AAO, 3-year treatment resistance, and clinical remission. We hypothesised that pathway-specific genetic loads drive distinct clinical trajectories, providing a mechanistic framework to stratify patients and identify individuals at high risk for early onset, clinical severity or primary treatment failure.

## Methods

### Sample

Clinical and genomic data were derived from two independent, well-characterised cohorts, comprising HC and patients diagnosed with SCZ-spectrum disorders according to the Diagnostic and Statistical Manual of Mental Disorders, Fourth Edition (DSM-IV) criteria [32]. Inclusion required a primary diagnosis of SCZ, or related psychotic spectrum disorders.

The Biomedical Research Network in Mental Health (CIBERSAM) cohort provided the primary large-scale dataset, comprising 3,856 individuals (2,145 patients from psychiatric in-patient units across seven medical centres and 1,711 HC) recruited in Spain [33]. This cohort was used to establish genomic associations with core clinical endophenotypes, specifically AAO and baseline symptom severity. Symptom severity was evaluated at study intake by certified research psychiatrists using the Positive and Negative Syndrome Scale (PANSS), capturing positive, negative, and general psychopathology subscales. AAO was defined as the age at which the first psychotic symptoms emerged, determined using a structured consensus protocol integrating reports from the treating psychiatrist, patient, and first-degree relatives. In cases where this information was not available, the date of the patient’s first psychiatric evaluation for psychotic symptoms was used [4]. Written informed consent was obtained from all participants (or legally authorised representatives) prior to enrolment, and research protocols were approved by the Clinical Research Ethics Committees of each participating hospital.

To evaluate the impact of genetic liability on long-term disease trajectories and treatment outcomes, we analysed the Cantabria Program for Early Intervention in Psychosis (PAFIP) cohort. PAFIP is a prospective longitudinal cohort that enrolled 739 individuals (552 patients and 187 HC) including patients presenting with a FEP between 2001 and 2018 [34]. FEP patients were assigned to three consecutive clinical trial phases (PAFIP I, II, and III), encompassing randomised, flexible-dose, and open-label clinical trials with a standardised 3-year prospective follow-up period. Antipsychotic dosing was adjusted at the treating psychiatrist’s discretion to maintain the minimum effective therapeutic dose, with medication switching permitted based on clinical efficacy and patient tolerability. Patients underwent longitudinal prospective evaluation at 3 years post-enrolment to assess i) treatment response and resistance using the Treatment Resistance and Response in Psychosis (TRRIP) consensus criteria [35]; and ii) long-term clinical remission of symptoms was defined according to the standardised criteria [36]. Sociodemographic characteristics were recorded at baseline. The research protocol was approved by the Clinical Research Ethics Committee of Cantabria, and all participants provided written informed consent.

### Genotyping, quality control, and imputation

Genome-wide array genotyping was conducted independently across both cohorts. The CIBERSAM cohort was array-genotyped using the Illumina Infinium PsychArray platform at the Broad Institute of MIT and Harvard as part of the Psychiatric Genomics Consortium (PGC) wave 3 SCZ GWAS meta-analysis [1]. The PAFIP cohort was array-genotyped using the Illumina Infinium Global Screening Array (GSA) v3.0. Quality control (QC) protocols were applied independently for each cohort prior to genotype imputation using PLINK v1.90b7.2 [37]. Briefly, QC included the sequential exclusion of samples and variants based on individual genotype missingness (>2%), sex discrepancies (reported males with *F* < 0.8; reported females with *F* > 0.2), minor allele frequency (MAF < 0.01), deviation from Hardy-Weinberg equilibrium (for HC *P* < 1e^−6^; for cases *P* < 1e^−10^), heterozygosity (mean (x□) ± 3 SD) and relatedness (pi-hat > 0.125). Finally, genotypes from each cohort were projected onto the 1000 Genomes Project Phase 3 reference panel (IGSR) using Principal Component Analysis (PCA). Genetically non-European individuals, defined as those exceeding 3 SD from the European population centroid along principal components 1 (PC1) and PC2, were excluded from downstream analyses. After QC, 3,044 individuals (1,755 patients and 1,289 HC) from the CIBERSAM cohort were included, of whom 1,544 (88%) patients had available AAO data and 440 (25%) had PANSS data. In the PAFIP cohort, 645 individuals (472 patients and 173 HC) were included in the analyses; among patients, 363 (77%) had 3-year follow-up data on symptom remission and 361 (76%) had treatment response data (Table 1).

**Table 1.**
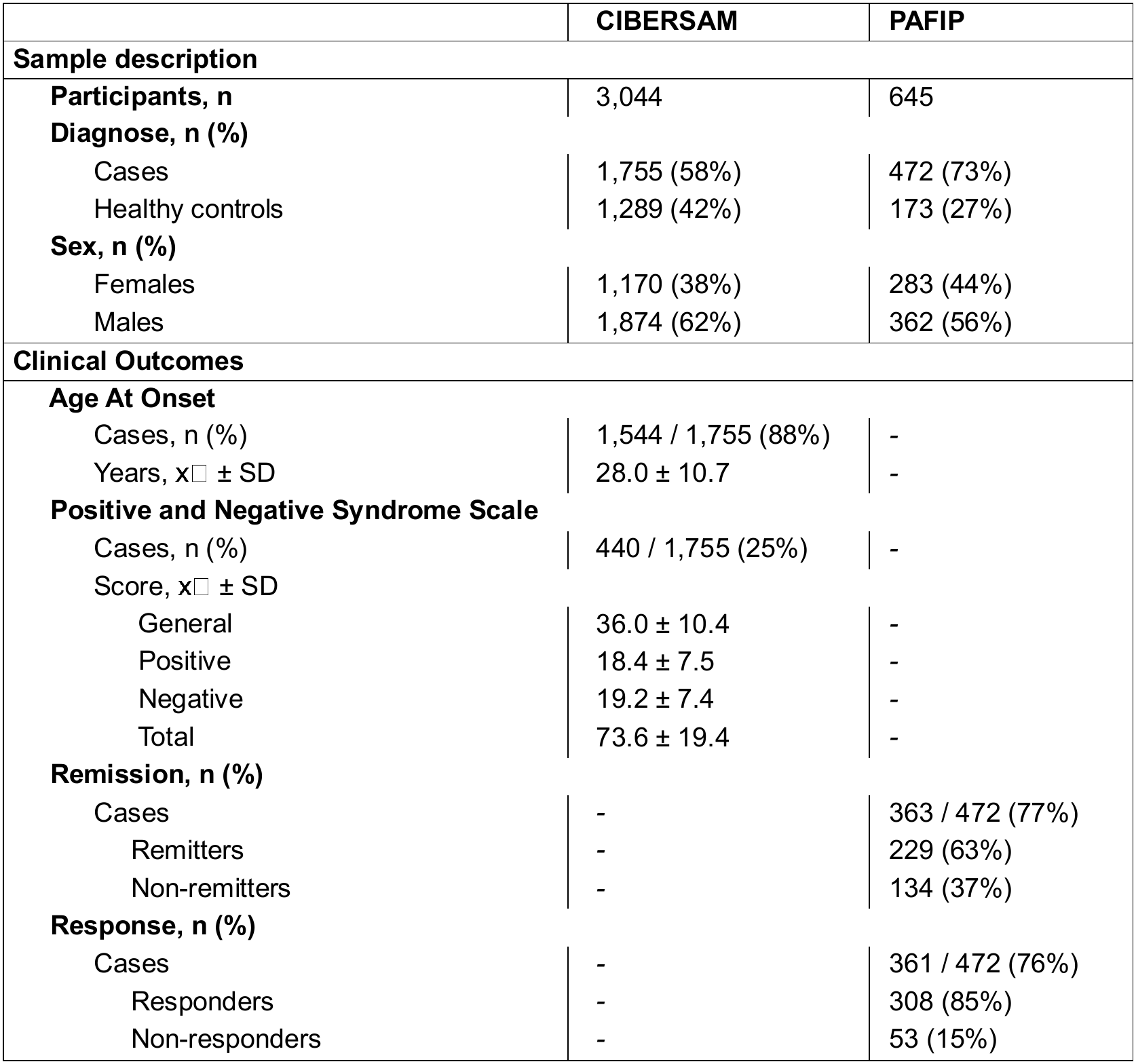
Cohort features after quality control. Number of individuals (n), percentage (%), mean (x□) and standard deviation (SD).

|  | **CIBERSAM** | **PAFIP** |
|---|---|---|
| **Sample description** |  |  |
| **Participants, n** | 3,044 | 645 |
| **Diagnose, n (%)** |  |  |
| Cases | 1,755 (58%) | 472 (73%) |
| Healthy controls | 1,289 (42%) | 173 (27%) |
| **Sex, n (%)** |  |  |
| Females | 1,170 (38%) | 283 (44%) |
| Males | 1,874 (62%) | 362 (56%) |
| **Clinical Outcomes** |  |  |
| **Age At Onset** |  |  |
| Cases, n (%) | 1,544 / 1,755 (88%) | - |
| Years, x□ ± SD | 28.0 ± 10.7 | - |
| **Positive and Negative Syndrome Scale** |  |  |
| Cases, n (%) | 440 / 1,755 (25%) | - |
| Score, x□ ± SD |  |  |
| General | 36.0 ± 10.4 | - |
| Positive | 18.4 ± 7.5 | - |
| Negative | 19.2 ± 7.4 | - |
| Total | 73.6 ± 19.4 | - |
| **Remission, n (%)** |  |  |
| Cases | - | 363 / 472 (77%) |
| Remitters | - | 229 (63%) |
| Non-remitters | - | 134 (37%) |
| **Response, n (%)** |  |  |
| Cases | - | 361 / 472 (76%) |
| Responders | - | 308 (85%) |
| Non-responders | - | 53 (15%) |

Genotype imputation was performed using the Michigan Imputation Server v2.0 platform [38]; accessed in November 2025) using the Haplotype Reference Consortium reference panel (HRC-r1.1). Genotypes were phased using EAGLE2 and subsequently imputed with minimac4. Post-imputation QC steps included removing variants with low imputation quality scores (r^2^ < 0.9 or INFO < 0.9), and low minor allele frequency (MAF < 0.001).

### Neurotransmitter pathway gene set curation and genomic annotation

To define genetic components of SCZ liability specific to individual NT pathways we systematically curated gene sets for five primary NT signalling networks: ACh, DA, GABA, Glu, and 5-HT. Biological pathways and ontologies were extracted from four comprehensive databases (accessed in May–June 2025): GeneOntology (GO) using AmiGO2, KEGG using KEGGREST (v1.48.0), Reactome using ReactomePA (v1.52.0), and WikiPathways using rWikiPathways (v1.28.1). Systematic query parameters were established for each NT system using specific inclusion and exclusion strings (ACh: inclusion of ach, acetylcholine, cholinergic, muscarinic, and nicotinic; exclusion of chromaffin, and gastric; DA: inclusion of dopa, and exclusion of dopachrome; GABA: inclusion of gaba, and gamma-aminobutyric acid; Glu: inclusion of glutamate, and glutamic acid; exclusion of taste, and tRNA; 5-HT: inclusion of serotonin, serotonergic, and 5-hydroxytryptamine). Genes were retained if captured in at least one database, yielding a comprehensive set of 1,478 non-redundant NT-associated genes: 302 for ACh, 322 for DA, 301 for GABA, 993 for Glu, and 210 for 5-HT (Figures 1 and S1; Table S1). Because broad glutamatergic ontologies capture ubiquitous metabolic processes distinct from central neurotransmission, the global Glu annotation inherently included pleiotropic genes involved in general cellular energy and amino acid metabolism. To isolate central synaptic signalling, we created a functionally refined Glu subset (Glu_Lite_; n = 912 genes). This refined set was confined to synaptic transmission, ionotropic/metabotropic receptor signalling, and vesicular transport machinery for parallel sensitivity analyses (see Supplementary Methods; Figure S1; Table S1).

**Figure 1.**
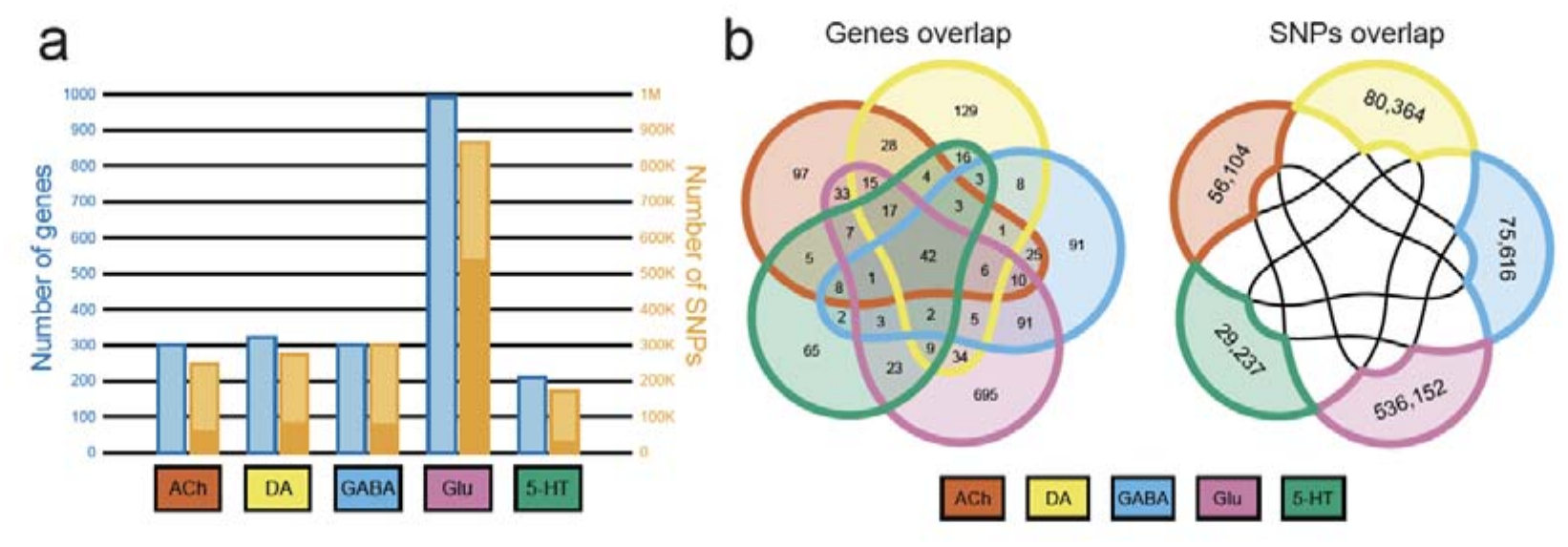
Identification and overlap of neurotransmitter-specific gene and variant sets. (a) Number of genes (in blue, left axis) and FUMA-annotated SNPs (in orange, right axis) assigned to each neurotransmitter pathway: acetylcholine (ACh), dopamine (DA), γ-aminobutyric acid (GABA), glutamate (Glu), and serotonin (5-HT). Dark orange represents specific SNPs related to each pathway. (b) Overlap genes and SNPs from neurotransmitter pathways.

Genomic coordinates for all curated target genes were mapped using the Ensembl database (GRCh37/hg19 assembly). To capture both proximal coding variation and distal cis-regulatory elements (enhancers/promoters), a 1Mb flanking genomic window was applied around gene boundaries. These intervals were intersected with SCZ GWAS summary statistics [1], yielding an initial pool of 4,273,261 SNPs. These SNPs were subsequently functionally annotated using Functional Mapping and Annotation of GWAS (FUMA v1.6.0; [39]. Variants were assigned to NT genes based on positional mapping, expression quantitative trait loci (eQTLs), and 3D chromatin interactions using a linkage disequilibrium (LD) independence threshold of r^2^ 0.8. This procedure yielded pathway-specific variant sets comprising 246,609 SNPs for ACh, 274,463 for DA, 301,221 for GABA, 864,587 for Glu (814,236 for Glu_Lite_), and 170,366 for 5-HT. Finally, to guarantee pathway independence and eliminate collinearity across polygenic scoring vectors, any SNP overlapping multiple systems within the set of 777,473 unique SNPs was excluded from the pathway-specific architectures. Thus, the final mutually exclusive pathway-specific variant sets comprised 56,104 SNPs for ACh, 80,364 for DA, 75,616 for GABA, 536,152 for Glu, and 29,237 for 5-HT (Figure 1; Table S2). Independence for each of the pathway-specific SNP sets was verified using LD (r^2^ ≥ 0.8), confirming their exclusivity (100% independence for ACh, 99.97% for DA, 99.98% for GABA, 99.98% for Glu, and 100% for 5-HT).

### Polygenic score (PGS) generation

We computed a gPGS for SCZ (gPGS_SCZ_) and pathway-specific sPGSs (sPGS_ACh_, sPGS_DA_, sPGS_GABA_, sPGS_Glu_, and sPGS_5-HT_) using PRSice-2 v2.3.5 [40], based on the European-ancestry SCZ GWAS summary statistics [1]. To prevent sample overlap, leave-one-out summary statistics excluding the CIBERSAM cohort were used. Following the framework established by [41], PGS were generated using a clumping– and-thresholding (C+T) approach across a grid of 100 parameter combinations. This matrix comprised 10 *P*-value thresholds (1, 0.5, 0.1, 5×10^-2^, 5×10^-3^, 5×10^-4^, 5×10^-5^, 5×10^-6^, 5×10^-7^, and 5×10^-8^) and 10 LD r^2^ clumping thresholds (1, 0.9, 0.8, 0.7, 0.6, 0.5, 0.4, 0.3, 0.2, and 0.1). To avoid the overfitting inherent in selecting a single optimal threshold, we performed PCA across the resulting 100-score matrix. For each participant, we kept the PC1, capturing the dominant shared variance across all combinations. In the CIBERSAM cohort, PC1 accounted for 80% of the variance for the gPGS and 61%, 69%, 66%, 68%, and 73% across the ACh, DA, GABA, Glu, and 5-HT sPGSs, respectively. Corresponding values in the PAFIP cohort were 85%, 57%, 72%, 64%, 67%, and 63%. Similar results for the PC1 variance were observed in the Glu_Lite_ sensitivity analysis (see Supplementary Methods). The resulting PC1 were regressed on sex and the first 10 ancestry PCs, and the resulting residuals were retained as the final adjusted PGS for all downstream analysis.

### Individual stratification and profiling

Individuals in the CIBERSAM cohort (both SCZ cases and HC) were stratified into deciles based on their gPGS_SCZ_ values to evaluate their case-control discriminatory capacity. Logistic regression models were used to estimate the relative odds of SCZ diagnosis across deciles. The 5^th^ decile was used as the reference for calculating Odds Ratios (ORs) with 95% Confidence Intervals (CIs). Global polygenic explanatory power was quantified in the continuous liability scale (r^2^_liability_) using the transformation proposed in [42], assuming a population prevalence (K) of 1% for SCZ spectrum disorders (Table S4).

To translate continuous polygenic distributions into categorical strata, participants (both patients and HC) were classified according to their NT-specific sPGS. Individuals with sPGS values in the highest decile (10^th^ decile) of a given NT pathway, were defined as a *High-Risk* group for that specific pathway. Subsequently, each individual’s top-decile status was mapped across all five NT systems. The profiles were named according to the corresponding NT-risk as *ACh_HR_*, *DA_HR_*, *GABA_HR_*, *Glu_HR_*, and *5-HT_HR_*. Individuals in the top decile for more than one NT pathway were excluded from the analyses. Individuals who did not rank in the top decile for any of the five evaluated NT-pathways constituted the Reference baseline group (Figure 2B and S2).

**Figure 2.**
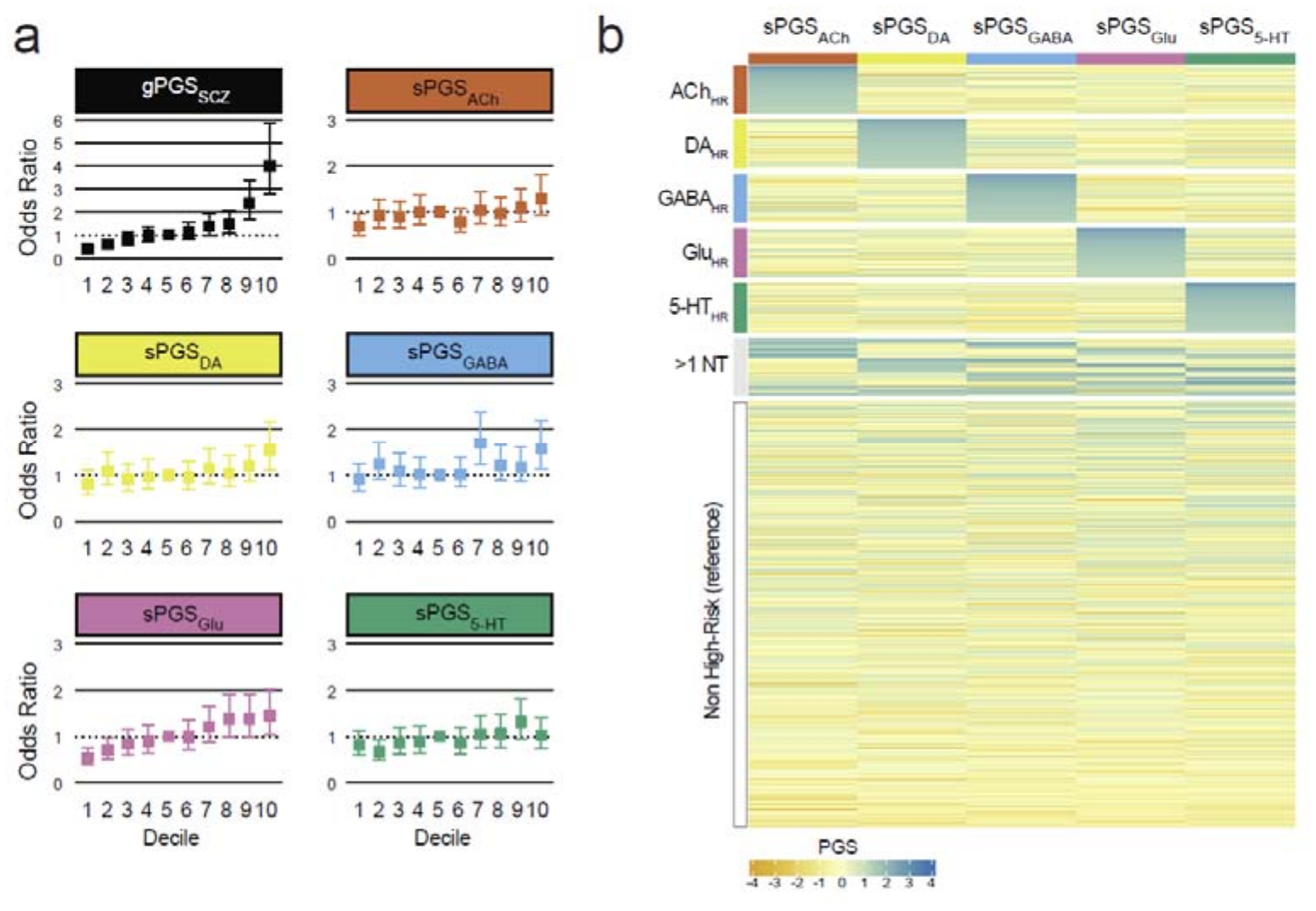
Distribution of polygenic scores and genetic stratification of CIBERSAM cohort. (a) Odds ratio for schizophrenia (SCZ) across polygenic scores (PGS) deciles of the genome-wide PGS for SCZ (gPGS_SCZ_) and each neurotransmitter (NT)-specific PGS (sPGS): acetylcholine (sPGS_ACh_), dopamine (sPGS_DA_), γ-aminobutyric acid (sPGS_GABA_), glutamate (sPGS_Glu_), and serotonin (sPGS_5-HT_). (b) Classification in *High-Risk* profiles according to sPGS across all participants. Individuals were classified based on the 10^th^ high-risk decile into a single NT *High-Risk* (*ACh_HR_*, *DA_HR_*, *GABA_HR_*, *Glu_HR_*, and *5-HT_HR_*), multiple NT *High-Risk* (>1 NT), or Non-High-Risk SPGs (*Reference*).

*High-risk* status was defined a priori as an sPGS on the 10^th^ decile to capture individuals at the extreme tail while retaining sufficient sample size for downstream comparisons. To assess whether the observed associations depended on this specific cutoff, complementary sensitivity analyses were performed employing less stringent decile thresholds to classify *High-Risk* and Reference individuals (see Supplementary Methods).

Although decile thresholds were established using the full cohort, downstream analyses were restricted to patients. Sex distribution across pathway risk profiles was evaluated using chi-square (χ²) tests, with statistical significance defined as *P* < 0.05. Continuous phenotypes — specifically AAO and PANSS subscale scores in the CIBERSAM cohort — were evaluated across genetic profiles using two-sided Wilcoxon rank-sum tests. Binary longitudinal outcomes, specifically 3-year treatment response and remission status in the PAFIP cohort, were evaluated using Fisher’s exact test. To control the type I error rate across multi-pathway hypothesis testing, all *P*-values across each studied phenotype were adjusted using the Benjamini-Hochberg False Discovery Rate (FDR) procedure. Statistical significance was defined as FDR < 0.05 and reported additionally to nominal *P*-values.

## Results

### Neurotransmitter-specific polygenic scores and participant stratification

We defined five NT-specific genetic architectures for ACh, DA, GABA, Glu, and 5-HT using curated gene sets and functional variant annotation. Variants shared across pathways were removed to generate mutually exclusive variant sets for downstream sPGS analyses (see Methods; Figure 1 and S1; Table S1).

To isolate system-specific signals from global liability, we generated a genome-wide polygenic score for SCZ (gPGS_SCZ_) and five pathway-stratified scores (sPGS) restricted exclusively to variants mapped within each curated NT gene set. Scores were residualised against biological sex and the top 10 genomic principal components (PCs) to control for population stratification in subsequent analyses.

The gPGS_SCZ_ robustly discriminated cases from HC (*P* = 2e^-16^), explaining 6.4% of the variance in SCZ liability. Among the NT-specific scores, sPGS_Glu_ showed the largest proportion of variance explained on the liability-scale (1.5%, *P* = 2.8e^-14^). This was followed by sPGS_DA_ (0.4%, *P* = 5.6e^-05^), sPGS_Ach_(0.3%, *P* = 3.9e^-04^), sPGS_GABA_ (0.3%, *P* = 1.2e^-03^) and sPGS_5-HT_ (0.3%, *P* = 7.3e^-04^). Because these scores were derived from non-overlapping variant sets, they may capture independent components of the overall SCZ polygenic liability (Table S4).

Stratification of individuals by gPGS deciles revealed a clear increase in the proportion of SCZ patients with increasing polygenic burden (Figure 2A and S2). Individuals in the highest gPGS_SCZ_ decile had approximately 4-fold higher odds of SCZ diagnosis than those in the 5^th^ decile (OR = 3.99, 95% CI = [2.76, 5.84], *P* = 4.24e^-10^; Table S5). Interestingly, stratifying in pathway-specific sPGS can also capture part of the SCZ case-control differentiation in the top decile, particularly for sPGS_DA_ (OR = 1.55, 95% CI = [1.12, 2.16], *P* = 9e^-3^), sPGS_GABA_ (OR = 1.58, 95% CI = [1.14, 2.19], *P* = 6e^-3^), and sPGS_Glu_ (OR = 1.45, 95% CI = [1.04, 2.01], *P* = 2.8e^-2^). However, membership in the highest decile of sPGS_ACh_ and sPGS_5-HT_ was not associated with SCZ diagnosis (ACh: OR = 1.29, 95% CI = [0.93, 1.80], *P* = 0.12; 5-HT: OR = 1.02, 95% CI = [0.74, 1.41], *P* = 0.91).

### NT-specific stratification identifies *high-risk* sub-architectures

To translate continuous polygenic variation into distinct genetic risk profiles, participants were stratified according to their sPGS values across the five NT pathways (see Methods). Application of this profiling framework to the CIBERSAM cohort (n = 3,044) revealed substantial heterogeneity in individual genetic profiles (Figure 2B; Table S6). Approximately one third of the cohort (33.2%, n = 1,010) was assigned to a single *High-Risk* profile, with proportions ranging from 6.4% to 6.7% across the five NT systems (n = 195 ACh, 204 DA, 204 GABA, 204 Glu, and 203 5-HT). An additional group of 237 individuals (7.8%) reached the top decile for two or more systems and were not considered for further analysis. The majority of the cohort (59%, n = 1,797) did not reach the top decile for any pathway and constituted the Reference group. Notably, these proportions (33.2%, 7.8%, 59.0%) closely matched those expected for five independent events at a 10% frequency (32.8%, 8.1%, and 59.0%, respectively).

A similar distribution of individuals was observed in the PAFIP cohort (n = 645; Table S6). Despite differences in clinical ascertainment and genotyping platform, the stratification proportions closely resembled those observed in CIBERSAM cohort. That is: 31.1% (n = 201) mapped to single-pathway *High-Risk* profiles, 8.4% (n = 54) to >1 *High-Risk* NT profile and excluded, and 60.5% (n = 390) to the *Reference* group. Re-profiling using the Glu_Lite_ definition produced similar category proportions: 91.6% of the individuals were retained their original profile (Table S6).

### Cholinergic and glutamatergic genetic profiles resolve clinical heterogeneity

To determine whether pathway-specific genetic liability was associated with distinct endophenotypes, we evaluated the relationship between sPGSs profiles and the clinical outcomes, including AAO, and PANSS symptom measures, in the CIBERSAM cohort. Sex distributions remained balanced across all pathway risk groups (*P* ≥ 0.05 for all comparisons; Table S7).

We first tested whether gPGS_SCZ_ was associated with clinical severity (Figure 3; Table S8). Individuals in the *High-Risk* group for SCZ (*SCZ_HR_*) showed no significant difference in AAO compared to the *Reference* group (x _SCZ_ = 28.1 vs. x _Reference_ = 28.0, 95% CI = [-1.00, 1.77], *P* = 0.708). Similarly, no significant differences were observed for PANSS total (x _SCZ_ = 74.5 vs. x _Reference_ = 72.4, 95% CI = [-6.00, 4.00], *P* = 0.749), general psychopathology (x _SCZ_ = 35.0 vs. x _Reference_ = 35.3, 95% CI = [-2.00, 4.00], *P* = 0.507), positive symptoms (x _SCZ_ = 19.2 vs. x _Reference_ = 18.3, 95% CI = [-3.00, 1.00], *P* = 0.409), and negative symptoms (x _SCZ_ = 20.3 vs. x _Reference_ = 18.8, 95% CI = [– 4.00, 3.32e-5], *P* = 0.12).

**Figure 3.**
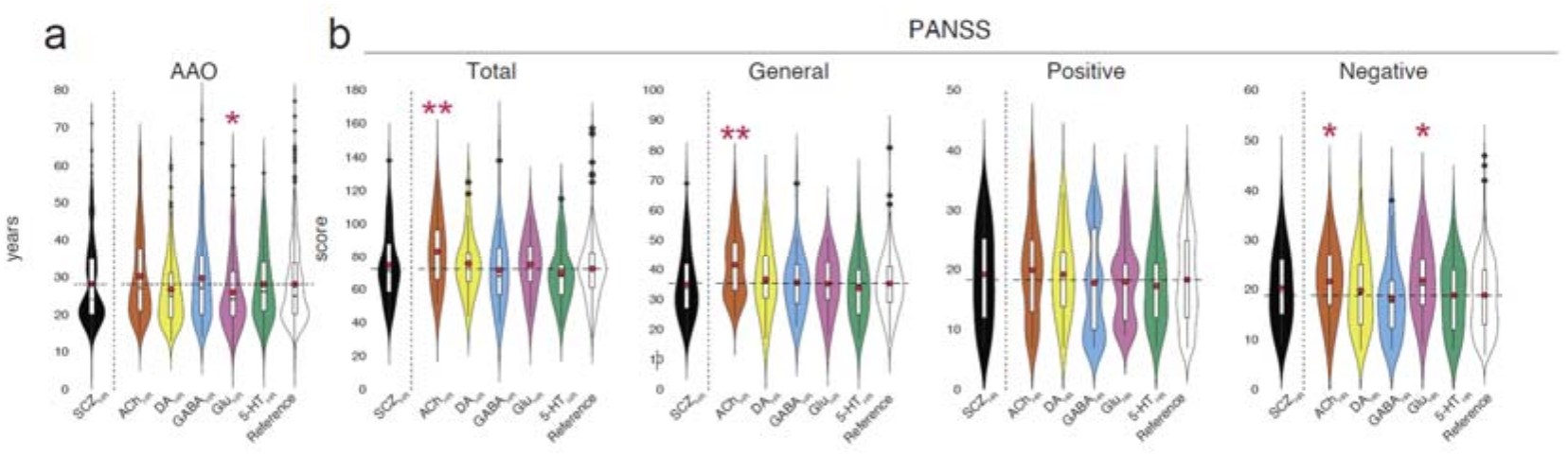
Effects of genetic *High-Risk* profiles on clinical phenotypes in the CIBERSAM cohort. (a) Age at onset (AAO) and (b) dimensions of the Positive and Negative Symptoms Scale (PANSS) across *High-Risk* genetic profiles of genome-wide for schizophrenia (*SCZ_HR_*) and neurotransmitter-specific: acetylcholine (*ACh_HR_*), dopamine (*DA_HR_*), γ-aminobutyric acid (*GABA_HR_*), glutamate (*Glu_HR_*), and serotonin (*5– HT_HR_*). The red dots indicate the group means whereas the dashed lines indicate the mean of the *Reference* profile. Differences were assessed using a two-sided Wilcoxon rank-sum test and significance is indicated by * (*P* < 0.05), and ** (FDR < 0.05).

In contrast, NT-specific stratification revealed pathway-specific differences in clinical phenotypes (Figure 3; Table S8). Individuals within the *Glu_HR_* profile showed a nominal association with AAO, developing their first psychotic symptoms on average 2.12 years earlier than the reference group (x _Glu_ = 25.9, 95% CI = [-3.35, –3.50e^-5^], *P* = 0.042, FDR = 0.188). Regarding symptomatology, individuals in the *ACh_HR_* showed higher overall symptom burden, specifically for PANSS total score (x _Glu_ = 83.1, 95% CI = [4.00, 18.00], *P* = 0.003, FDR = 0.017) and the general psychopathology subscale score (x _Glu_ = 41.6, 95% CI = [2.00, 10.00], *P* = 0.002, FDR = 0.012). Nominal associations with higher negative symptom scores were also observed for the *ACh_HR_* and *Glu_HR_* profiles (x _ACh_ = 21.6, 95% CI = [1.00, 6.00], *P* = 0.019, FDR = 0.71; and x _Glu_ = 21.9, 95% CI = [4.07e-5, 6.00], *P* = 0.028, FDR = 0.71, respectively). No significant differences were observed for the PANSS positive subscale across any of the NT-specific profiles. Consistent patterns of association were observed across alternative decile thresholds and using the Glu_Lite_ definition, indicating that the main findings were not restricted to the primary classification method (Figure S3A).

### Glutamatergic and GABAergic genetic profiles associate with longitudinal treatment outcomes

To evaluate whether NT-specific profiles were associated with long-term treatment outcomes, we analysed 3-year symptomatic remission and treatment response within the prospective PAFIP cohort. Sex distributions did not differ significantly between pathway risk groups for any comparison (*P* ≥ 0.05 for all comparisons; Table S7).

First, no significant association was observed between gPGS and either remission (*P =* 0.299; Table 2) or treatment response (*P* = 0.803; Table 3). In contrast, NT-specific profiles revealed selective associations with treatment outcomes (Table 2 and 3). Patients in the *Glu_HR_* profile exhibited the most severe clinical trajectories, with 3.66-fold higher odds of clinical non-remission at 3 years compared with the reference group (OR = 3.66, 95% CI = [1.26, 11.51], *P* = 9e^-03^, FDR = 0.049). The *Glu_HR_* profile also showed a nominal association with treatment non-response (OR = 3.37, 95% CI = [0.96, 10.61], *P* = 0.03, FDR = 0.14). Similarly, the *GABA_HR_* profile exhibited a nominal association with non-remission (OR = 2.72, 95% CI = [1.08, 7.01], *P* = 0.02, FDR = 0.06), whereas no association was observed with treatment non-response (*P* ≥ 0.05). The remaining profiles — *ACh_HR_*, *DA_HR_*, and *5-HT_HR_*— showed no significant association with either non-remission or treatment non-response (*P* ≥ 0.05).

**Table 2.**
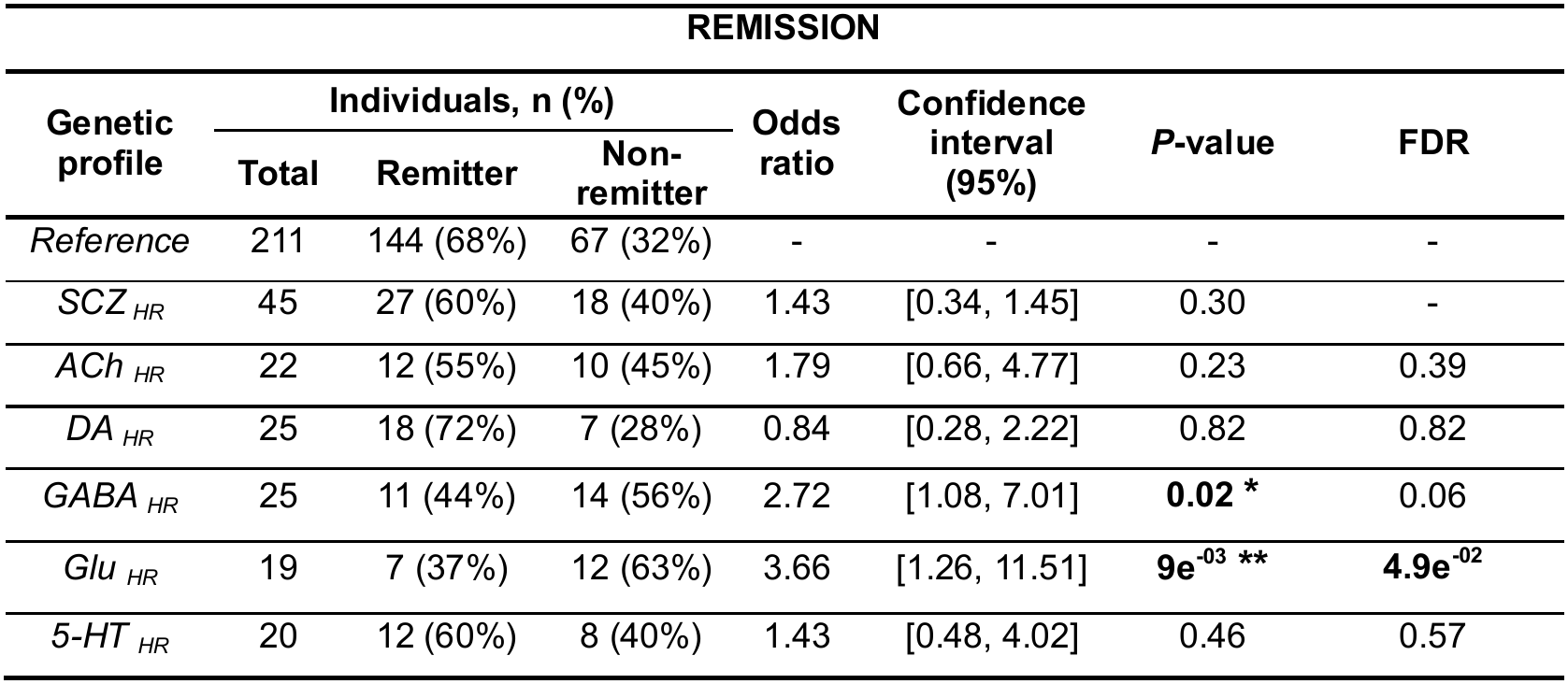
Association of *High-Risk* genetic profiles with 3-year follow-up remission of symptoms in the PAFIP cohort. Profile differences were assessed using a Fisher’s exact test against *Reference* group, with non-remitters serving as the reference category. *High-Risk* profiles: schizophrenia (*SCZ_HR_*), acetylcholine (*ACh_HR_*), dopamine (DA*_HR_*), γ-aminobutyric acid (*GABA_HR_*), glutamate (*Glu_HR_*) and serotonin (*5-HT_HR_*). Statistically significant results are shown in bold. Number of individuals (n), percentage (%). Significance is indicated by * (*P* < 0.05), ** (FDR < 0.05).

| **REMISSION** |  |  |  |  |  |  |  |
|---|---|---|---|---|---|---|---|
| **Genetic profile** | **Individuals, n (%)** |  |  | **Odds ratio** | **Confidence interval (95%)** | ***P*-value** | **FDR** |
|  | **Total** | **Remitter** | **Non-remitter** |  |  |  |  |
| *Reference* | 211 | 144 (68%) | 67 (32%) | - | - | - | - |
| *SCZ* <sub>*HR*</sub> | 45 | 27 (60%) | 18 (40%) | 1.43 | [0.34, 1.45] | 0.30 | - |
| *ACh* <sub>*HR*</sub> | 22 | 12 (55%) | 10 (45%) | 1.79 | [0.66, 4.77] | 0.23 | 0.39 |
| *DA* <sub>*HR*</sub> | 25 | 18 (72%) | 7 (28%) | 0.84 | [0.28, 2.22] | 0.82 | 0.82 |
| *GABA* <sub>*HR*</sub> | 25 | 11 (44%) | 14 (56%) | 2.72 | [1.08, 7.01] | **0.02 \*** | 0.06 |
| *Glu* <sub>*HR*</sub> | 19 | 7 (37%) | 12 (63%) | 3.66 | [1.26, 11.51] | **9e<sup>-03</sup> \*\*** | **4.9e<sup>-02</sup>** |
| *5-HT* <sub>*HR*</sub> | 20 | 12 (60%) | 8 (40%) | 1.43 | [0.48, 4.02] | 0.46 | 0.57 |

**Table 3.**
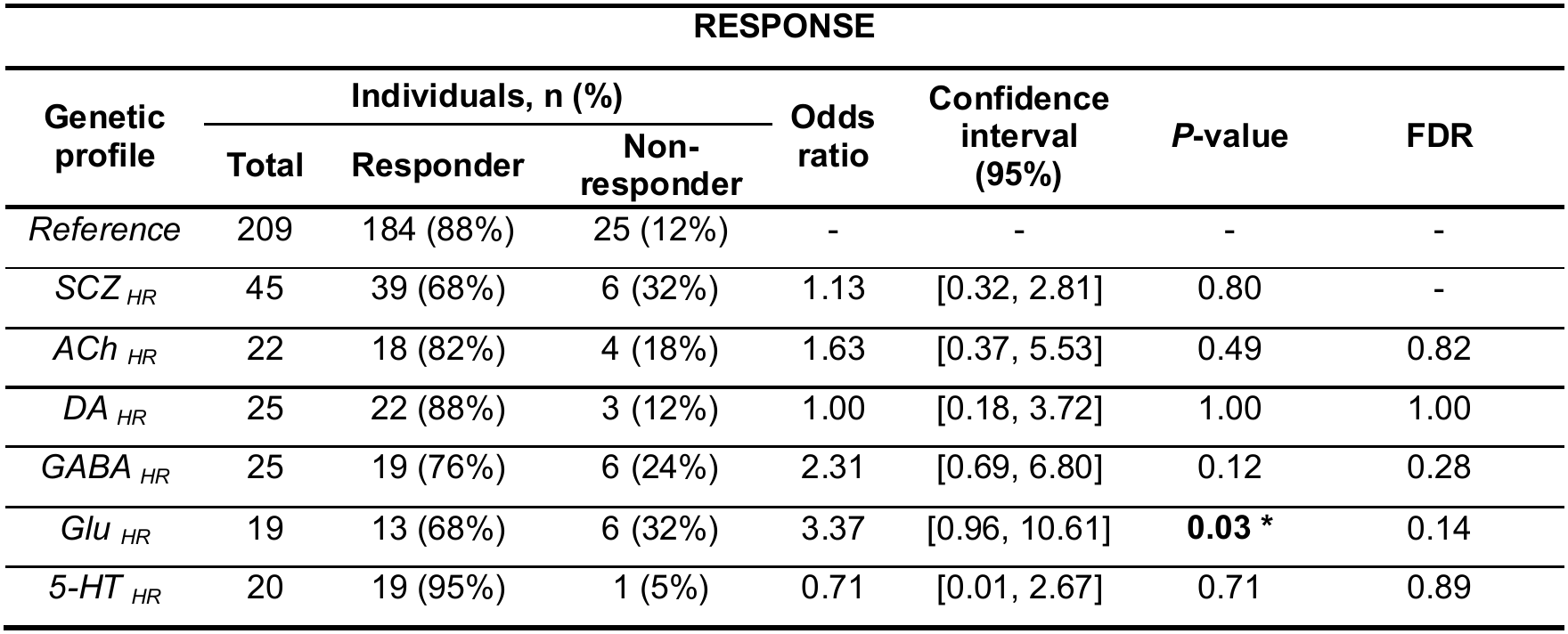
Association of *High-Risk* genetic profiles with 3-year follow-up response to treatment in the PAFIP cohort. Profile differences were assessed using a Fisher’s exact test against *Reference* group, with non-responders serving as the reference category. *High-Risk* profiles: schizophrenia (*SCZ_HR_*), acetylcholine (*ACh_HR_*), dopamine (*DA_HR_*), γ-aminobutyric acid (*GABA_HR_*), glutamate (*Glu_HR_*) and serotonin (*5-HT_HR_*). Statistically significant results are shown in bold. Number of individuals (n), percentage (%). Significance is indicated by * (*P* < 0.05).

| RESPONSE |  |  |  |  |  |  |  |
|---|---|---|---|---|---|---|---|
| **Genetic profile** | **Individuals, n (%)** |  |  | **Odds ratio** | **Confidence interval (95%)** | ***P*-value** | **FDR** |
|  | **Total** | **Responder** | **Non-responder** |  |  |  |  |
| *Reference* | 209 | 184 (88%) | 25 (12%) | - | - | - | - |
| *SCZ* <sub>*HR*</sub> | 45 | 39 (68%) | 6 (32%) | 1.13 | [0.32, 2.81] | 0.80 | - |
| *ACh* <sub>*HR*</sub> | 22 | 18 (82%) | 4 (18%) | 1.63 | [0.37, 5.53] | 0.49 | 0.82 |
| *DA* <sub>*HR*</sub> | 25 | 22 (88%) | 3 (12%) | 1.00 | [0.18, 3.72] | 1.00 | 1.00 |
| *GABA* <sub>*HR*</sub> | 25 | 19 (76%) | 6 (24%) | 2.31 | [0.69, 6.80] | 0.12 | 0.28 |
| *Glu* <sub>*HR*</sub> | 19 | 13 (68%) | 6 (32%) | 3.37 | [0.96, 10.61] | **0.03 \*** | 0.14 |
| *5-HT* <sub>*HR*</sub> | 20 | 19 (95%) | 1 (5%) | 0.71 | [0.01, 2.67] | 0.71 | 0.89 |

Finally, the patterns of association with longitudinal treatment outcomes were broadly preserved when using the Glu_Lite_ variant definition and across alternative decile cutoffs, further supporting the consistency of the observed effects (Figure S3B; see Supplementary Methods).

## Discussion

Understanding the profound clinical and biological heterogeneity of SCZ remains one of the challenges in modern psychiatric genomics [43]. Although GWAS have established the highly polygenic architecture of SCZ [1], standard gPGS_SCZ_ reduce information from thousands of loci into a single value. While effective for population-level diagnostic case-control discrimination, they have limited ability to resolve variation in clinical phenotypes among affected individuals. In this study, we partitioned SCZ genetic liability into system-specific sPGS spanning five primary NT pathways (ACh, 5-HT, DA, GABA, and Glu) in two independent cohorts. This revealed distinct associations with AAO, symptom dimensions, and longitudinal treatment outcomes, supporting the use of biologically informed genetic stratification as a potential framework for dissecting clinical heterogeneity in SCZ.

Regarding clinical symptomatology, the *ACh_HR_* profile was associated with greater overall and general psychopathology, while both *ACh_HR_*and *Glu_HR_* showed nominal associations with greater negative symptom burden. This is clinically relevant because negative symptoms are among the most difficult symptom domains to treat effectively with current antipsychotic therapies [48, 49]. The association between *ACh_HR_*risk and psychopathology is consistent with the central role of muscarinic and nicotinic signalling in the cognitive and negative symptom domains of SCZ [21]. Disruption of cholinergic transmission can impair auditory sensory gating, attentional filtering, and motivational processing [50, 51]. This genetic architecture is also consistent with the therapeutic efficacy of dual M_1_/M_4_ muscarinic receptor agonists, such as xanomeline– trospium (KarXT), which have been shown to significantly reduce PANSS symptom scores without inducing extrapyramidal motor symptoms or metabolic disruption [52]. Regarding Glu, in FEP patients, levels of Glu are correlated with PANSS scores [53], and have been reported to induce both positive symptoms and cognitive deficits [24]. In contrast, positive psychotic symptoms showed no significant differences across NT-specific profiles in our study. This finding supports the hypothesis that hallucinations and delusions may represent convergent downstream manifestations of network dysfunction arising from upstream genetic mechanisms [17, 54].

Standard first-line antipsychotics primarily exert their therapeutic efficacy primarily through dopamine D2 receptor antagonism [55]. It has consequently been hypothesised that the ∼30% of SCZ patients who are treatment-resistant represent a distinct “normodopaminergic” subtype driven by upstream cortical dysfunctions rather than primary striatal dopamine hyperactivity [54, 56]. Our findings are consistent with this hypothesis, suggesting that both GABAergic and glutamatergic genetic profiles are linked to an increased likelihood of persistent symptoms over the 3-year follow-up period. While global polygenic liability was not associated with 3-year treatment outcomes, the *Glu_HR_* profile was associated with an increased likelihood of non-remission. There were also additional nominal associations between *Glu_HR_* and treatment non-response, as well as between *GABA_HR_*liability and non-remission. Consistent with this hypothesis, *DA_HR_* showed no association with treatment resistance. This suggests that, when disease liability is driven by lesions in the canonical dopaminergic pathway, the resulting clinical phenotype may be highly responsive to standard D2 blockade. However, when the aetiology is rooted in Glu or GABA signalling networks, downstream D2 antagonism is pharmacologically insufficient, manifesting clinically as treatment-resistant SCZ. Our findings, also align with neuroimaging data demonstrating that treatment-resistant patients lack the elevated striatal dopamine synthesis capacity observed in treatment-responsive patients [18, 57]. Recent studies highlight disruptions in glutamatergic neurotransmission — particularly N-methyl-D-aspartate (NMDA) receptor hypofunction— as well as GABAergic interneuron signalling [1, 58]. Additionally, rare variant studies have implicated genes involved in synaptic pruning and NMDA receptor subunits function in cognitive impairment and treatment resistance [16, 59, 60].

AAO demonstrates substantial additive heritability [4]. However, studies have either failed to detect robust associations or have only detected small variances in global polygenic burden, possibly due to signal dilution through non-developmental pathways [4, 44, 45]. By shifting the analytic focus, our results revealed that genetic liability predisposition concentrated within the glutamatergic pathway (*Glu_HR_*) significantly accelerates disease manifestation, bringing forward the onset of first psychotic symptoms by more than two years. Glutamatergic signalling is essential for cortical maturation, synaptic plasticity, and synaptic pruning during adolescent neurodevelopment [46, 47]. Consequently, a high genetic burden within these excitatory networks likely disrupts normal synaptic refinement, potentially contributing to earlier disease onset. This pathway-specific association is consistent with recent large-scale and functional genomic analyses demonstrating that SCZ risk variants are highly enriched in excitatory pyramidal neurons active during adolescent remodelling, [1, 11].

There are several methodological limitations that warrant further consideration. First, to analyse the specific effects of NT and avoid multicollinearity, we excluded pleiotropic SNPs shared by multiple NT systems. While necessary to isolate pathway-specific main effects, it inherently discards ‘hub’ genes that are likely to contribute to complex psychiatric phenotypes. Second, as the NT-specific risk architectures were derived from GWAS summary statistics of European ancestry, the trans-ethnic portability of these genetic architectures is unknown. Third, our models lack granular data on antipsychotic medication, dosage, and cumulative exposure. As these treatments can cause significant changes to NT networks, there is a potential risk of confounding factors. This highlights the need for well-characterised cohorts with detailed medication exposure data in future investigations. Finally, due to sample size constraints, we were unable to perform fully powered sex-stratified analyses. Given well-established sex differences in SCZ age at onset, clinical presentation, and disease trajectories, larger multi-site cohorts will be required to determine whether pathway-specific risk architectures exhibit sex-dependent differential prognostic utility.

In conclusion, this study supports the potential value of biologically informed polygenic stratification for investigating clinical heterogeneity in SCZ. While global polygenic burden fails to differentiate symptom profiles, stratifying this monolithic risk into NT-specific sub-architectures reveals association with certain outcomes in SCZ. Specifically, genetic liability within the glutamatergic, GABAergic and cholinergic pathways is associated with an earlier disease onset, greater psychopathology, and an increased risk of treatment non-remission. NT-specific polygenic scores may be particularly valuable in the early stages of psychosis by identifying distinct genetic risk profiles associated with symptom severity and long-term clinical outcomes, potentially enabling earlier risk stratification and more personalised therapeutic approaches.

## Acknowledgements

This work was supported by project 202230 from the Fundació La Marató de TV3 to GS and GM, and by the Catalan Agency of Research and Universities (AGAUR, grant number 2021SGR01065) to EV. GS is also supported by the Agencia Estatal de Investigación under grants PID2025-173577NB-I00 and PID2022-140137NB-I00, and by the Instituto de Salud Carlos III with co-financing by European Funds (under grant CP20/00064 for the Miguel Servet Contract to GS; and PI24/01118 to GM).

## Data availability statement

The GWAS summary statistics used to construct the PGS are publicly available via the PGC data portal. We used the leave-one-out version of these statistics. Individual-level clinical phenotypic data and raw human genomic data generated for both cohorts are not publicly available due to ethical restrictions but can be obtained from the authors upon reasonable request.

## Code Availability Statement

All the bioinformatic pipelines, and figure-processing workflows generated during this study are open source and publicly archived on GitHub at https://github.com/gap-irbcatsud-huipm/Neurotransmitters_PGSs.

## Author contributions

PR-C performed all the analyses. GM and GS conceived the study and designed the experimental and analytical framework. PR-C prepared all figures and wrote the manuscript draft. GM and GS reviewed and refined the final manuscript. JV-B, BC-F, MDM, MJE, JG-P, CA, AG-P, LF, BA, JB, JC, LM, and EV provided the clinical and genomic data. All authors critically reviewed, revised, and approved the final manuscript for publication.

