## Supplementary Methods for "Neurotransmitter-specific polygenic scores dissect clinical heterogeneity in schizophrenia"

### **Glu_Lite_ parallel analysis**

In parallel, a reduced version of the Glu pathway (Glu_Lite_) was evaluated. To exclude genes primarily associated with metabolic processes while retaining those more directly involved in synaptic signalling, a second filtering step was applied to the gene-group names obtained from the ontologies. Gene groups containing the terms *receptor*, *synapse*, *synaptic*, *nerve*, *uptake*, or *secretion* were retained. This resulted in 912 genes across the four biological databases, corresponding to 92% of the initial gene set (Figure S1; Table S1). Thus, taking into account the other NT sets, a total of 1,400 unique genes were used for this parallel analysis.

Accordingly, among the set of 738,677 unique SNPs in the Glu_Lite_ parallel study, the exclusive pathway-specific variant sets used to calculate the sPGS comprised 56,844 SNPs for ACh, 81,256 for DA, 78,826 for GABA, 492,020 for Glu_Lite_, and 29,731 for 5-HT (Table S3).

Individuals from the CIBERSAM and PAFIP cohorts were then stratified based on the sPGS calculated from these variant sets, following the same procedure described in the main Methods. The results for the PC1 variance in ACh, DA, GABA, Glu, and 5-HT sPGSs were 61%, 69%, 66%, 68%, and 73% respectively in the CIBERSAM cohort, and 85%, 57%, 72%, 63%, 67%, and 63% in the PAFIP cohort. As shown, these results are the same as for the main approach results, except for the GABA in PAFIP. The analysis yielded stratifying patterns is similar to those obtained using the standard approach (Table S6).

### **PGS high genetic risk thresholds**

To stratify individuals within each cohort according to their genetic risk, predefined PGS thresholds were used to classify individuals' risk into *High-Risk* or *Non-High-Risk* groups.

In addition to the 10^th^ decile (>90% of the PGS population distribution) used to define the *High-Risk* group, less stringent thresholds were also evaluated. Specifically, individuals in the 9^th^ decile or above (>80%) were classified as *High-Risk* under an alternative definition.

The two *High-Risk* definitions were combined with different *Non-High-Risk* thresholds, resulting in five risk-stratification approaches: (i) three approaches used the 10^th^ decile (>90%) as the *High-Risk* threshold, combined with *Non-High-Risk* thresholds ≤9, ≤8, and ≤7 deciles (≤90%, ≤80%, and ≤70%, respectively; showed in the Figure S3 as 10_9, 10_8, and 10_7, respectively); and (ii) two approaches used the 9^th^ decile or above (>80%) as the *High-Risk* threshold, combined with *Non-High-Risk* thresholds of ≤8 and ≤7 deciles (≤80% and ≤70%, respectively; showed in the Figure S3 as 9_8, and 9_7, respectively).
