## Supplementary Figures for "Neurotransmitter-specific polygenic scores dissect clinical heterogeneity in schizophrenia"

**Figure S1.** **Neurotransmitter-related genes identified from biological databases.** Number of genes (n) associated with the five neurotransmitter pathways – acetylcholine (ACh), dopamine (DA), γ-aminobutyric acid (GABA), glutamate (Glu), and serotonin (5-HT) – derived from the four biological databases – GeneOntology, KEGG, Reactome, and WikiPathways –. The refined Glu subset (Glu_Lite_) is also shown.

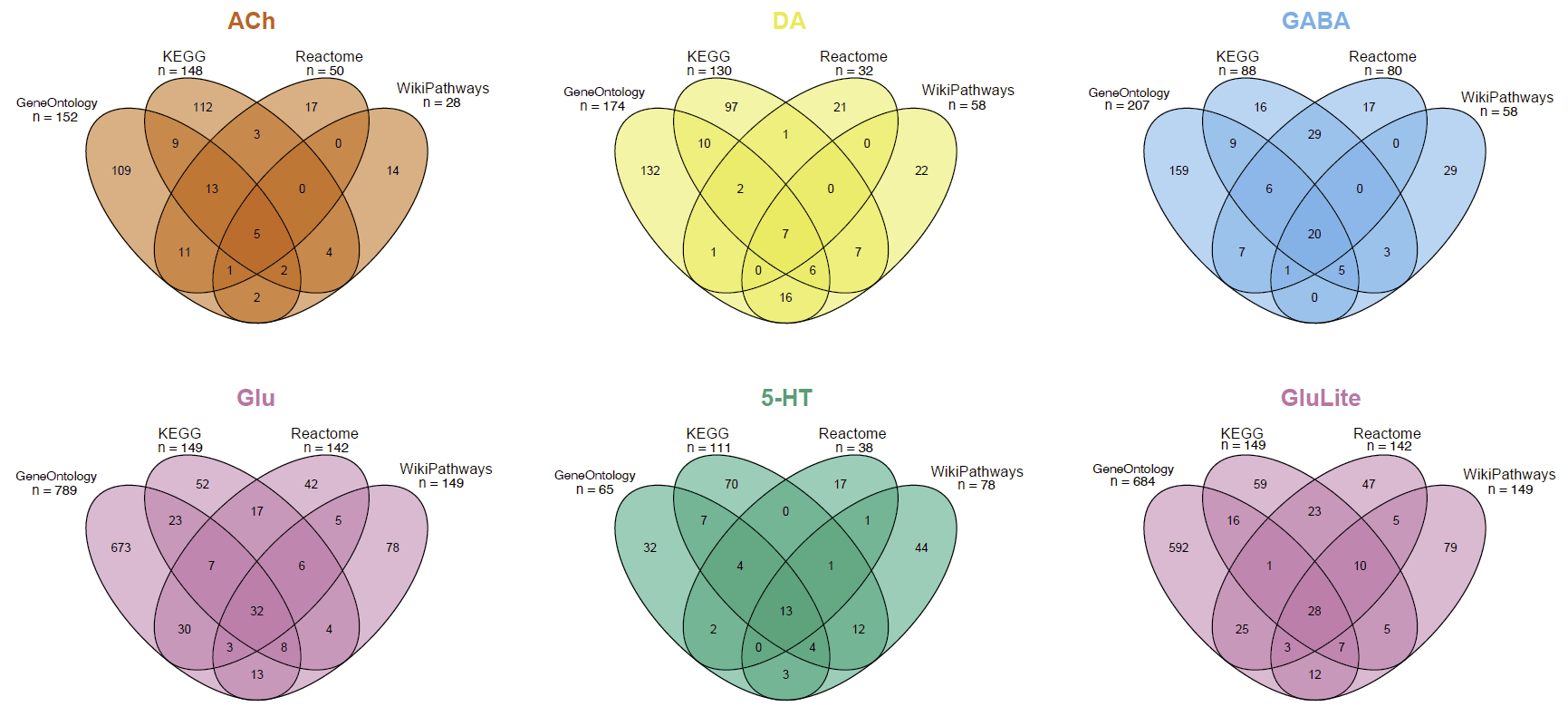

**Figure S2.** **Distribution of CIBERSAM individuals according to decile-based stratification.** The proportion of patients (in orange) and healthy controls (in blue) is shown across population deciles (1 to 10) based on polygenic scores (PGS): the genome-wide for schizophrenia (gPGS_SCZ_) and each neurotransmitter-specific – acetylcholine (sPGS_ACh_), dopamine (sPGS_DA_), γ-aminobutyric acid (sPGS_GABA_), glutamate (sPGS_Glu_), and serotonin (sPGS_5-HT_) –.

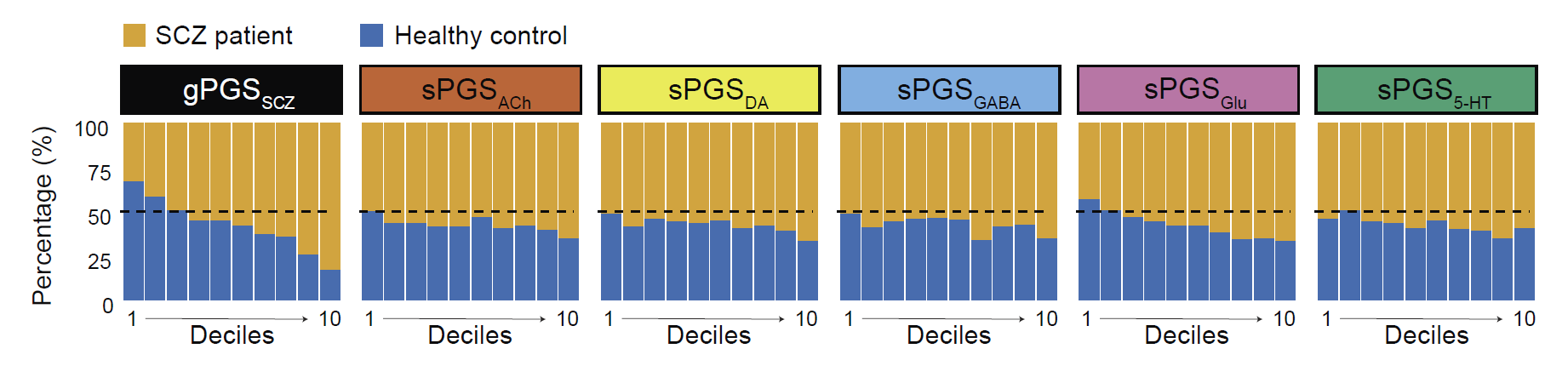

**Figure S3.** **Associations between neurotransmitter-related *High-Risk* profiles and clinical outcomes across multiple genetic risk thresholds.** Effects on clinical outcomes across *High-Risk* genetic profiles of genome-wide for schizophrenia (*SCZ_HR_*) and neurotransmitter-specific –acetylcholine (*ACh_HR_*), dopamine (*DA_HR_*), γ-aminobutyric acid (*GABA_HR_*), glutamate (*Glu_HR_*), and serotonin (*5-HT_HR_*)–. (a) Associations with age at onset (AAO) and Positive and Negative Syndrome Scale (PANSS) subscales scores in the CIBERSAM cohort. (b) Associations with treatment response and remission at 3 years in the PAFIP cohort. Predefined genetic risk thresholds were used to classify individuals into *High-Risk* or *Non-High-Risk* profiles (see Supplementary Methods). Highlighted red cells indicate the principal condition used in main figures. Cell colour indicates the direction of the association (higher values in blue and lower values in orange) relative to the *Reference* profile. Darker shading of the cells indicates lower *P*-values, whereas lighter shading indicates non-significant associations. Significance is indicated by · (*P* < 0.1), * (*P* < 0.05), ** (*P* < 0.01), and *** (*P* < 0.001).

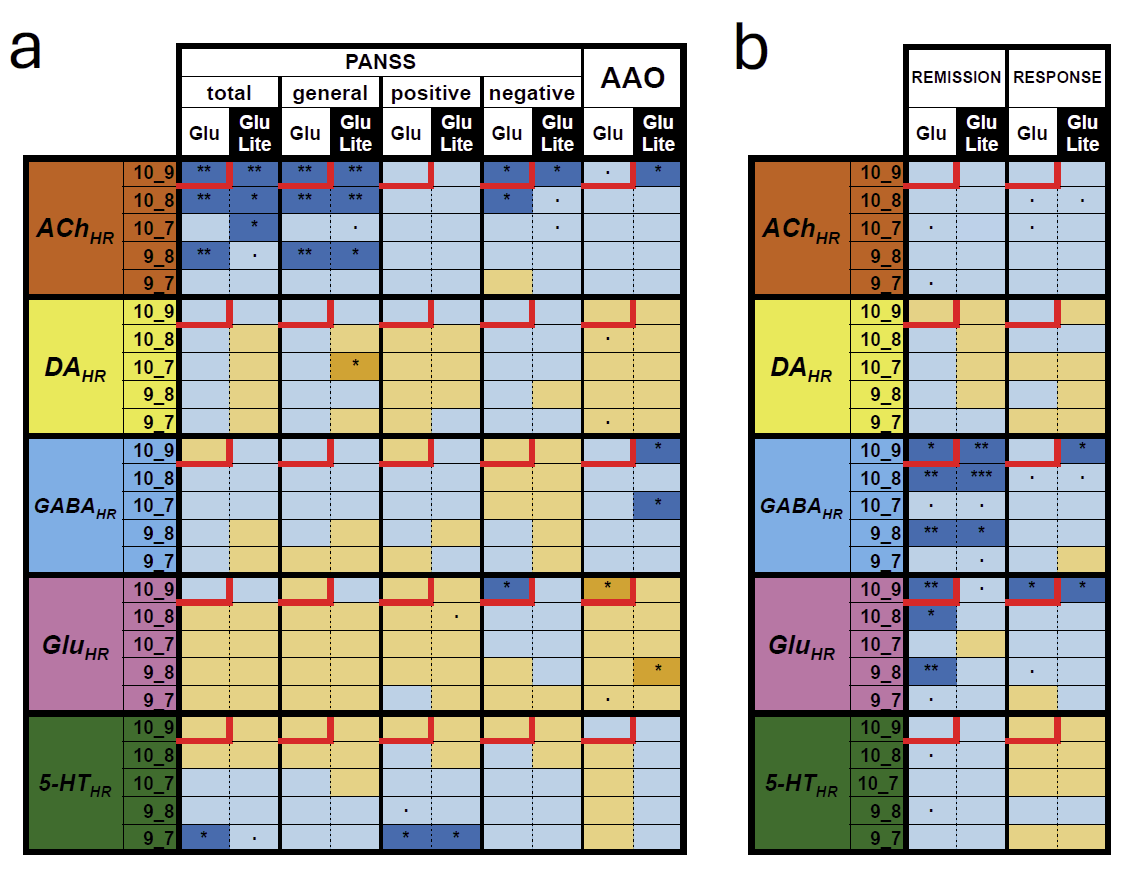
